# Common X-chromosome variants are associated with Parkinson’s disease risk

**DOI:** 10.1101/2020.12.18.20248459

**Authors:** Yann Le Guen, Valerio Napolioni, Michael E. Belloy, Eric Yu, Lynne Krohn, Jennifer A. Ruskey, Ziv Gan-Or, Gabriel Kennedy, Sarah J. Eger, Michael D. Greicius

## Abstract

**Objective:** Identify genetic variants on the X-chromosome associated with Parkinson’s disease (PD) risk.

**Methods:** We performed an X-chromosome-wide association study (XWAS) of PD risk by meta-analyzing results from sex-stratified analyses. To avoid spurious associations, we designed a specific harmonization pipeline for the X-chromosome and focused on a European ancestry sample. We included 11,324 cases, 280,060 controls, and 5,379 proxy cases, based on parental history of PD. Additionally, we tested the association of significant variants with: (i) PD risk in an independent replication with 1,564 cases and 2,467 controls, and (ii) putamen volume in 33,360 individuals from the UK Biobank.

**Results:** In the discovery meta-analysis, we identified: rs7066890 (OR=1.10 [1.06-1.14]; P=2.2×10^−9^) intron of *GPM6B*, and rs28602900 (OR=1.10 [1.07-1.14]; P=1.6×10^−8^) in a high gene density region including *RPL10, ATP6A1, FAM50A, PLXNA3*. The rs28602900 association with PD was replicated (OR=1.16 [1.03-1.30]; P=0.016) and shown to colocalize with a significant expression quantitative locus (eQTL) regulating *RPL10* expression in the putamen and other brain tissues in GTEx. Additionally, the rs28602900 locus was found to be associated with reduced brain putamen volume. No results reached genome-wide significance in the sex-stratified analyses.

**Interpretation:** We report the first XWAS of PD and identify two genome-wide significant loci. The rs28602900 association replicated in an independent PD dataset and showed concordant effects in its association with putamen volume. Critically, rs26802900 is a significant eQTL of *RPL10*.These results support a role for ribosomal proteins in PD pathogenesis and show that the X-chromosome contributes to PD genetic risk.

## Introduction

Parkinson’s Disease (PD), a neurodegenerative disorder characterized by a broad range of motor and non-motor symptoms, is thought to be caused by a combination of aging, genetics and environmental factors. PD affects the nervous system at multiple levels from the enteric nervous system to the cortex, but prominent motor symptoms associated with PD are linked to the substantia nigra and striatum. PD heritability has been estimated at 34% in a twin study ^1^ and 16 to 36% in genome-wide association studies (GWAS) ^2^. The latter estimates were derived from autosomal variants and did not include the genetic contribution of sex chromosomes. Similarly, the latest PD GWAS meta-analysis, which identified 90 common genetic variants associated with PD risk, focused solely on autosomes ^2^. The effect of common sex chromosome variants on PD risk remains unexplored, despite the relative risk for PD being 1.5 times greater in males than in females ^3–6^. Various mechanisms have been proposed to account for this sex difference in PD risk, including sex hormone levels and other environmental and genetic factors influenced by biological sex ^7,8^. To date, a few studies have identified variants or loci on the X-chromosome linked to PD, notably, a linkage analysis in PD multiplex families which identified the *PARK12* locus ^9^. However this locus is quite large and has not yet been linked to a gene ^10^. Whole-exome sequencing studies in pedigree and single case reports have linked loss of function mutations on *RAB39B* with Waisman syndrome, an X-linked early-onset, α-synucleinopathy with Parkinsonism and intellectual disability ^11–14^.

Though the X-chromosome is 155 Mb and accounts for 5% of the human genome, it remains excluded from the vast majority of GWAS because it requires different quality control than autosomal chromosomes ^15^ due to the way it is inherited. Females inherit one X-chromosome from each parent, while males only inherit a maternal copy and outside of pseudoautomosal regions, male X-chromosomes do not undergo any recombination during meiosis. This causes the X-chromosome to have reduced genetic diversity. Notably, the X-chromosome structure is more sensitive to historical events such as population bottleneck, sex-biased demographics events, and has a different mutation rates than autosomes ^16^. Additionally, until recently, genotyping arrays were poorly designed for the X-chromosome ^15^.

Our work addresses this gap by conducting the first PD X-chromosome-wide association study (XWAS). Specifically, we performed meta-analyses including all publicly available PD cohorts with X-chromosome data, as well as the UK Biobank cohort, in which we used both diagnosed PD individuals and proxy-cases based on parental history of PD. The use of this PD proxy phenotype was recently shown to provide increased power for the discovery of PD genetic risk factors ^2^. We first meta-analyzed males and females separately to identify potential sex specific effects. Then, to increase statistical power, we meta-analyzed across the sexes to identify variants influencing PD risk in a sex-independent fashion. Variants that reached genome-wide significance were tested for replication in an independent PD meta-analysis and tested for association with putamen volume in the UK Biobank, and functionally annotated in gene expression datasets through colocalization analyses, leading to the identification of a putatively causal gene for PD that may provide novel targets for future therapeutic development.

## Methods

### Samples

For the discovery PD meta-analysis, we used two types of samples: PD cohorts published in previous studies and the UK Biobank, using both directly diagnosed PD and a proxy phenotype based on parental history of PD. The PD cohorts include the International PD Genomics Consortium (IPDGC) NeuroX dataset ^17^, the National Institute of Neurological Disorders and Stroke (NINDS) PD dataset ^18^, the Autopsy-Confirmed PD GWAS Consortium (APDGC) dataset genotyped by the Center for Inherited Disease Research (CIDR), the NeuroGenetics Research Consortium (NGRC) dataset ^19^, the UK PD Consortium and The Wellcome Trust Case Control Consortium 2 ^20^, referred to here as EBI MERGE. The whole-genome sequencing (WGS) data were obtained from the Accelerating Medicines Partnership - Parkinson’s Disease (AMP-PD), composed of the following cohorts: New Discovery of Biomarkers (BioFIND), the Harvard Biomarker Study (HBS), the Parkinson’s Progression Markers Initiative (PPMI) ^21^, and the Parkinson’s Disease Biomarkers Program (PDBP). **Table 1** shows a detailed description of the cohorts including genotyping platform, number of X-chromosome single nucleotide polymorphisms (SNPs) and demographics.

**Table 1.**
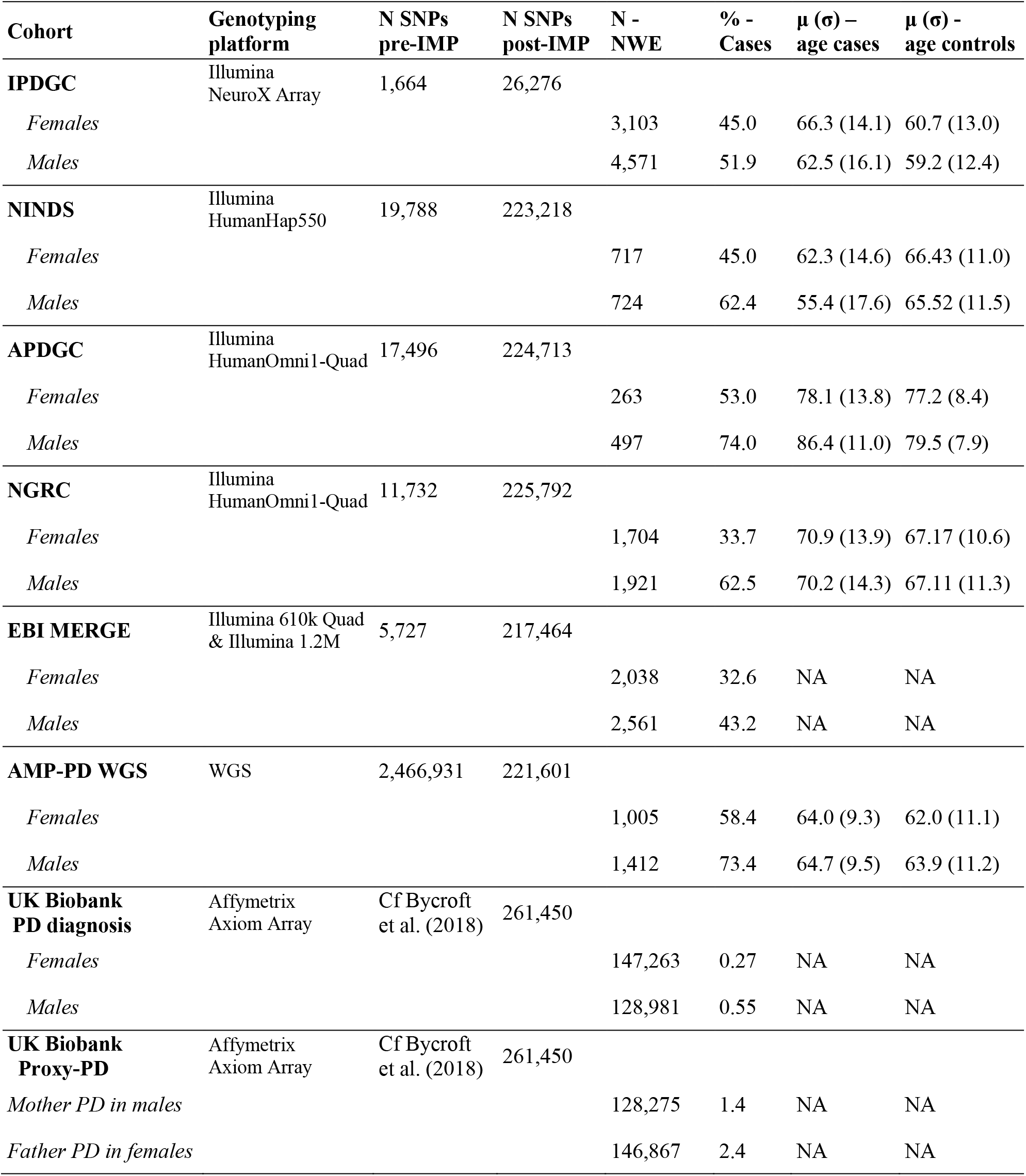
Overview of the cohorts included in the Parkinson’s disease discovery meta-analysis. *N SNPs pre-IMP*: number of SNPs passing quality control prior to imputation, or number of SNPs in the WGS. *N SNPs post-IMP*: number of SNPs with imputation quality r^2^ > 0.3 (N) and after filtering for MAF > 0.01 & genotyping rate > 95%. NWE: Northwestern Europeans.

In the replication, we meta-analyzed two PD datasets (**Table 2**): the first composed of the Parkinson’s Disease Cognitive Genetics Consortium (PDCGC) ^22^ and healthy individuals from the Adult Change in Thought (ACT) longitudinal cohort ^23^, referred to here as PDCGC & ACT, and the second from McGill University (McGill) including samples from the Quebec Parkinson Network (QPN)^24^.

**Table 2.** Overview of the cohorts included in the Parkinson’s disease replication meta-analysis. *N SNPs pre-IMP*: number of SNPs passing quality control prior to imputation, or number of SNPs in the WGS. *N SNPs post-IMP*: number of SNPs with imputation quality r^2^ > 0.3 (N) and after filtering for MAF > 0.01 & genotyping rate > 95%. NWE: Northwestern Europeans.

| Cohort | Genotyping platform | N SNPs pre-IMP | N SNPs post-IMP | N - NWE | % - Cases |
| --- | --- | --- | --- | --- | --- |
| **PDCGC & ACT** | Illumina NeuroX & Illumina 660W | 1,322 in PDCGC<br>8,908 in ACT | 29,283 in both |  |  |
| *Females* |  |  |  | 1,099 | 20.2 |
| *Males* |  |  |  | 1,141 | 36.5 |
| **McGill** | Illumina Infinium OmniExpress-24 | 15,912 | 227,144 |  |  |
| *Females* |  |  |  | 758 | 41.7 |
| *Males* |  |  |  | 1,033 | 59.1 |

The putamen volume analysis consisted of individuals with magnetic resonance imaging (MRI) scans that were successfully processed by the UK Biobank FSL pipeline ^25^ as of January 2020. The putamen volume used in our analysis corresponds to the average of the left and right hemispheres. UK Biobank details regarding the MRI scan parameters, and image quality control are described elsewhere ^26,27^.

The current study protocol was granted an exemption by the Stanford University institutional review board because the analyses were carried out on deidentified, off-the-shelf data; therefore, further informed consent was not required.

### Quality control, ancestry determination and imputation

To perform the first PD XWAS, we introduced a quality control and harmonization pipeline of X-chromosome SNP-array data from homogenous ancestry samples prior to X imputation. Autosomal variants were extracted from the SNP array genetic data and processed through our harmonization pipeline in several stages. First, we removed multi-allelic SNPs, SNPs located on structural variants and duplicated or monomorphic SNPs. The list of multi-allelic SNPs or SNPs located on common structural variants was created using Tri-Typer ^28^ and gnomAD ^29^. Additionally, we removed variants located on potential probe polymorphisms identified from gnomAD. These are defined as SNPs for which the probe may have variable affinity due to the presence of other SNP(s) within 20 bp and with minor allele frequency (MAF) above 1%. SNPs were checked for consistency with the Haplotype reference consortium (HRC) panel ^30^ using a checking script developed by the McCarthy group (well.ox.ac.uk/∼wrayner/tools/). This script enabled us to flip SNPs reported on the incorrect strand and excluded palindromic SNPs and SNPs with more than 10% MAF difference from the reference panel.

Using these quality control SNPs as inputs, we further excluded SNPs with a genotyping rate < 95% and Hardy-Weinberg equilibrium (HWE) p < 10^−5^ in controls and also removed individuals with more than 5% genotype missingness. Next, the individual ancestry was determined with SNPweights v.2.1 ^31^ using reference populations from the 1000 Genomes Consortium ^32^. By applying an ancestry percentage cut-off > 75%, the samples were stratified into the five super populations, South-Asians, East-Asians, Americans, Africans and Europeans. Since most of the samples belonged to the European population, we also determined their percent ancestry to three major ethnicities, Northwestern, Southeastern, and Ashkenazi Jewish, using reference populations available from SNPweights v.2.1. European subjects were stratified into the above-mentioned ethnicities when their ancestry percentage was greater than 50%. Further analyses were focused on the Northwestern European (NWE) subsample, which represents the vast majority of subjects and provided a highly homogenous sample. Two main reasons explain the more extreme population structure on the X-chromosome compared to autosomes: (i) the X-chromosome has a smaller effective population size and thus the rate of genetic drift of X-linked loci is amplified, (ii) local adaptation will lead to a higher levels of differentiation between geographically isolated population ^33^. As such, to better control our population structure, we decided to be stringent and restrict our analyses to NWE subjects.

Additionally, we merged all genotype quality control autosomes and performed an identity-by-descent on the whole NWE sample to identify duplicates and first-degree relatives. If the diagnosis was incongruent between cohorts for a duplicate subject, we chose the cohort with a PD diagnosis. Otherwise, we kept the subject’s data in the cohort with the following order of priority based on sequencing coverage of the X chromosome: AMP-AD WGS, APDGC, EBI MERGE, NINDS, NGRC, and IPDGC. Similarly, for first degree relatives, given the direction of the sex bias in PD risk, we prioritized the inclusion of a male relative over a female one. Additionally, we kept cases over controls, used the same order of priority for cohort selection when relatives had the same diagnosis and chose the older of the two relatives if they were controls in the same cohort.

The X chromosome variants underwent a similar harmonization pipeline as the autosomes. We excluded multi-allelic SNPs, SNPs within structural variations, and potential probe polymorphism SNPs. Additionally, our analysis excluded the pseudo autosomal regions of the X chromosome and used only the NWE subsample as derived above. Several steps were performed to avoid spurious findings: (1) variants with less than 95% genotyping rate and (2) individuals with more than 5% genotype missingness were excluded. (3) Reported sex was checked using PLINK1.9 ^34^ *--check-sex* flag, with 0.4 max value for females and 0.94 min value for men and all individuals with a discordant sex label were excluded. (4) Heterozygous SNPs in males were set as missing in males, while (5) SNPs with differential missingness between PD cases and controls were removed (p < 10^−5^). (6) HWE was tested in female controls and SNPs with p < 10^−5^ were removed. (7) Any monomorphic SNPs that remained were removed. (8) Differential missingness and differential MAF between males and females were both tested and SNPs with p < 10^−5^, for either one of the tests, were excluded. Finally, as for the autosomes, (9) remaining SNPs were checked for consistency with the HRC panel, flipping palindromic SNPs and excluding SNPs with more than 10% MAF difference from the reference panel.

The EBI MERGE dataset, which includes two SNP arrays, had these quality control steps performed separately on each array and the remaining SNPs were merged and re-processed through the whole harmonization pipeline. Similarly, PDCGC & ACT autosomes were first harmonized separately, then merged and re-harmonized together before ancestry determination and PCA computation. By contrast, the X-chromosome data were imputed separately and merged after imputation, due to a low X coverage of the NeuroX array used in PDCGC that resulted in only 111 overlapping ACT-PDCGC variants after harmonization.

The remaining X chromosome quality controlled SNPs were imputed on the Michigan Imputation Server (v1.2.4) ^35^, which uses minimac 4 for imputation and improved X chromosome support. The following parameters were selected: reference panel HRC r1.1 2016 (hg19) with the EUR subpanel, phasing Eagle v2.4, r-square imputation score cut off 0.3. Before the XWAS with PD status, SNPs with MAF < 0.01 or genotyping rate < 95% were removed in WGS and imputed data. Genetic principal components (PC) were computed within the NWE subsample for each cohort to account for population stratification in the downstream analysis ^36^. PCs were inspected for each cohort and indicated a homogenous sample.

The UK Biobank genotyping, imputation and quality control data are described in detail elsewhere 37. We considered the 33,360 British individuals with MRI scan, whose genetic data successfully passed the UK Biobank quality control and were not diagnosed with PD, nor had a parent with PD.

### Study-level analyses and meta-analysis

To identify sex-specific effects, we first performed sex-stratified meta-analyses separating males and females. We then performed a combined meta-analysis across all sex-stratified cohorts, to identify sex-independent effects with increased statistical power.

The male meta-analysis included 5,745 cases and 4,875 controls of NWE ancestry from PD cohorts, 706 PD cases and 128,275 controls from the UK Biobank unrelated British males, and 3,551 proxy male cases and 143,316 controls using the father’s PD status in unrelated British females. The rationale is that the female dosage, in this case, corresponds to the probability of the father carrying the variant and testing it against his PD status. Put differently, if a female has two copies of the effect allele, then her father is a carrier; if a female has one copy then there is 50% chance that her father is a carrier, if a female has no copies then her father is a non-carrier. The female meta-analysis included 3,104 cases and 4,925 controls of NWE ancestry from PD cohorts, 396 PD cases and 146,867 controls from the UK Biobank unrelated British females, and 1,828 proxy female cases and 126,447 controls using the mother’s PD status in unrelated British males. In this scenario, the mother must carry at least one copy for a male carrier and zero or no more than one copy for a male non-carrier. We excluded directly diagnosed PD subjects from the proxy phenotype analysis.

The associations were estimated with PLINKv2.0 ^34^ using the *–glm* flag, which performs a standard logistic regression for case/control phenotype. For each cohort, we covaried by the age provided in each study and the 10 PCs computed within the NWE sample. In each separate study, cases and controls were roughly age matched, with cases slightly older than controls (**Table 1**). This small age difference per cohort resulted in a small positive effect of age on PD risk that was adjusted for, except in the EBI MERGE cohort where age was unavailable. The age provided varied across studies between age at onset, age at clinical diagnosis or age at death for cases, and age at last exam or age at death for controls. To address this heterogeneity, we ran the analysis with and without age as a covariate and the loci passing the suggestive threshold did not change. For the UK Biobank, the covariates used were the first 10 PCs provided by the UK Biobank and the age at last visit of the participant. Finally, we meta-analyzed all XWAS summary statistics using fixed effect meta-analysis as implemented in GWAMA ^38^. Additionally, GWAMA tests for sex-heterogenous effects by combining male and female effects into a chi-squared distribution with one degree of freedom (assuming the same allelic effect in males and females) and testing the null hypothesis that the association is the same in males and females. The genome-wide significance threshold was set at p < 5×10^−8^ and the suggestive significance threshold at p < 10^−5^, corresponding to X-chromosome-wide correction.

We queried the expression quantitative loci (eQTL) among our XWAS loci in brain tissues of the Genotype-Tissue Expression (GTEx) v8 ^39^. GTEx (gtexportal.org) includes bulk RNASeq data from 12 brain tissues types: amydgala (n = 129), anterior cingulate cortex (n = 147), caudate (n = 194), cerebellar hemisphere (n = 175), cerebellum (n = 209), cortex (n = 205), frontal cortex (n = 175), hippocampus (n = 165), hypothalamus (n = 170), nucleus accumbens (n = 202), putamen (n = 170), and substantia nigra (n = 114). Additionally, we queried the Braineac database ^40^ (braineac.org), which includes n = 134 individuals with up to 12 different brain regions sequenced with RNA exon arrays. The eQTLs nominal significance, as well as false discovery rate (FDR) correction^41^, were performed by GTEx ^39^ and Braineac ^40^. Colocalization was performed using the R package *coloc* ^42^ and we report the posterior probability of colocalization (PP4) between PD and QTL associations in **Figures S1-S4**. Colocalization between XWA and eQTL results were inspected with *locuscompareR* ^43^ for all FDR significant results from GTEx.

We tested the association of the significant loci with putamen volume using a Bayesian linear mixed model as implemented in LMM-BOLT ^44^. In this second validation analysis males and females were analyzed together coding males as 0/2 and females as 0/1/2. We adjusted the analysis by brain volume, age at MRI scan, genotyping array, sex, and 10 PCs accounting for population stratification.

## Results

### Parkinson’s Disease X-chromosome discovery meta-analyses

We meta-analyzed all XWAS summary results in males, females, and in both sexes combined (**Fig 1**). In visually inspecting each cohort’s QQ plots and genomic control lambdas, we observed no inflation (**λ** = 0.983-1.042). As a sensitivity analysis, we also conducted the analysis without accounting for any covariates. The top associations remained unchanged and lambdas remained within the normal range (**λ** = 0.971-1.069).

**Figure 1.**
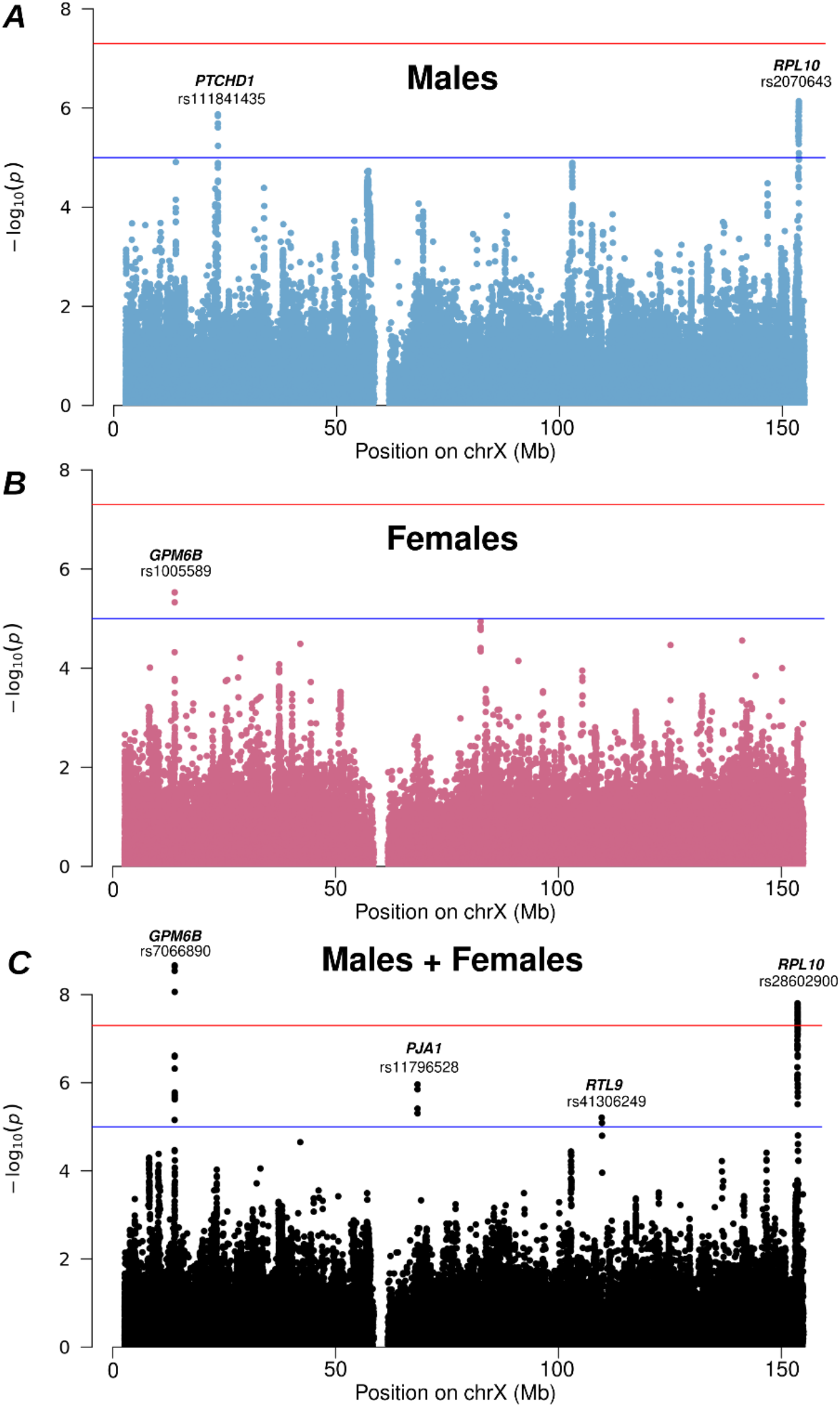
X-chromosome wide analyses reveal two genome-wide significant loci in the combined sex meta-analysis. X-chromosome wide sex-stratified discovery analyses in males **(A)** and in females **(B)**, and combined sex meta-analysis **(C)**. The most significantly regulated gene in brain tissues (*RPL10)* or the nearest gene (*GPM6B, PJA1, PTCHD1, RTL9*) is annotated on top of the lead SNP at each locus. The red line is the genome-wide significance threshold (p < 5×10^−8^) and the blue line is the suggestive significance threshold (p < 10^−5^), corresponding to X-chromosome-wide correction.

In the combined sex meta-analysis (**λ** = 1.041), two loci reached genome-wide significance with top SNPs: rs7066890 near *GPM6B* (OR = 1.10, MAF = 0.16, p = 2.2×10^−9^), rs28602900 near *RPL10* (OR = 1.10, MAF = 0.12, p = 1.6×10^−8^). For each SNP, the minor allele was associated with increased risk and in all analyses described below the rsID will refer to the minor allele. These two SNPs had a concordant direction of effect across cohorts (**Fig 2**). The first SNP is an intron of *GPM6B*, and the second SNP is located in a region with a high gene density (39 genes, within +/- 400kb).

**Figure 2.**
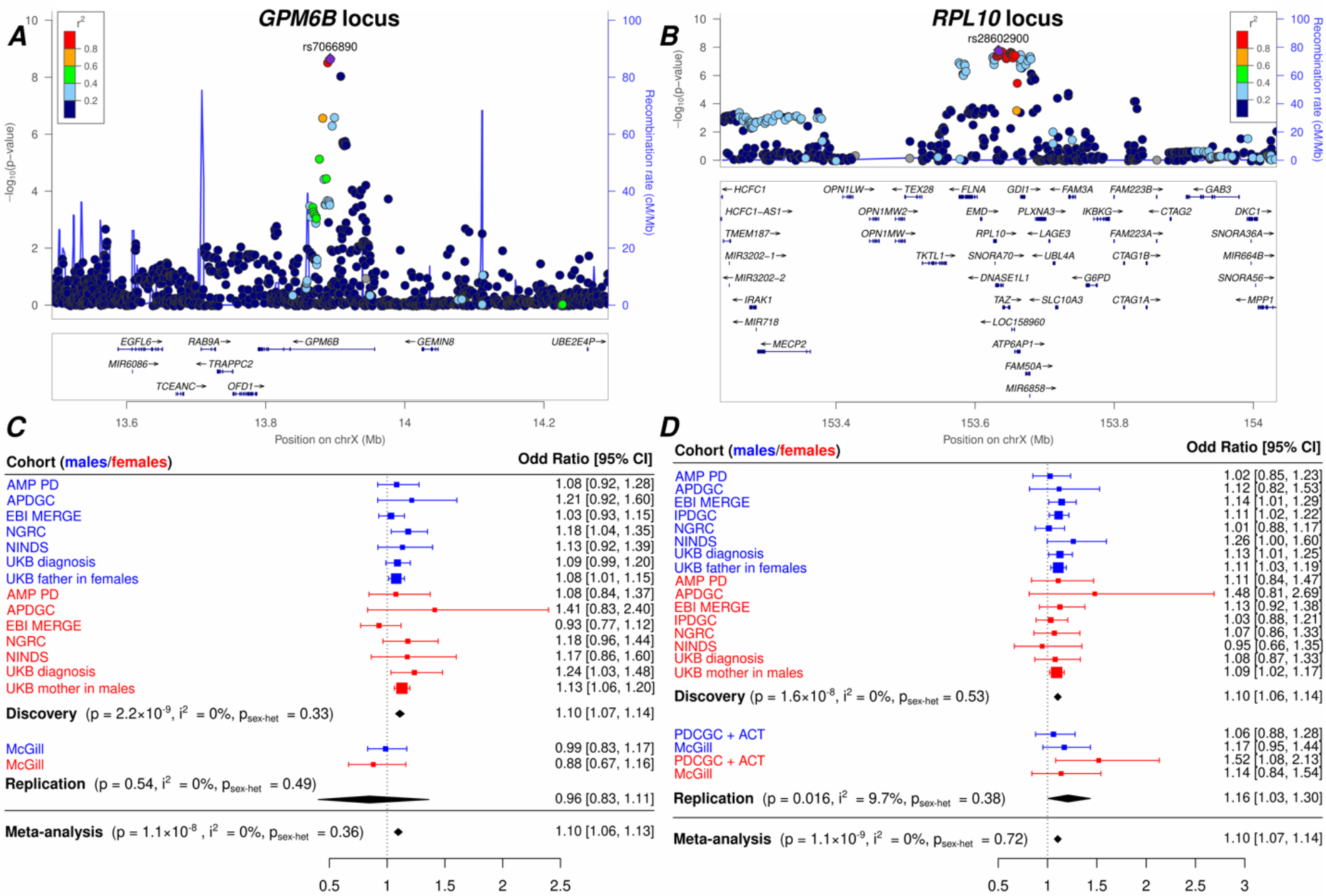
Regional details and forest plots of the genome-wide significant loci in the combined sex meta-analysis. Locuszoom plots of the *GPM6B* **(A)** and *RPL10* **(B)** loci, and the forest plots for their respective lead variant emphasizing their consistent direction of effect across datasets **(C, D)**.

For the sex-stratified analyses (**λ**_**male**_ = 1.004, **λ**_**female**_ = 0.980) there were no genome-wide significant results. Loci passing the suggestive, X-chromosome-wide significance threshold are shown in **Figure 1** and detailed in **Table 3**. The two genome-wide significant loci from the combined sex meta-analysis had a non-differentiated effect across sex.

**Table 3.**
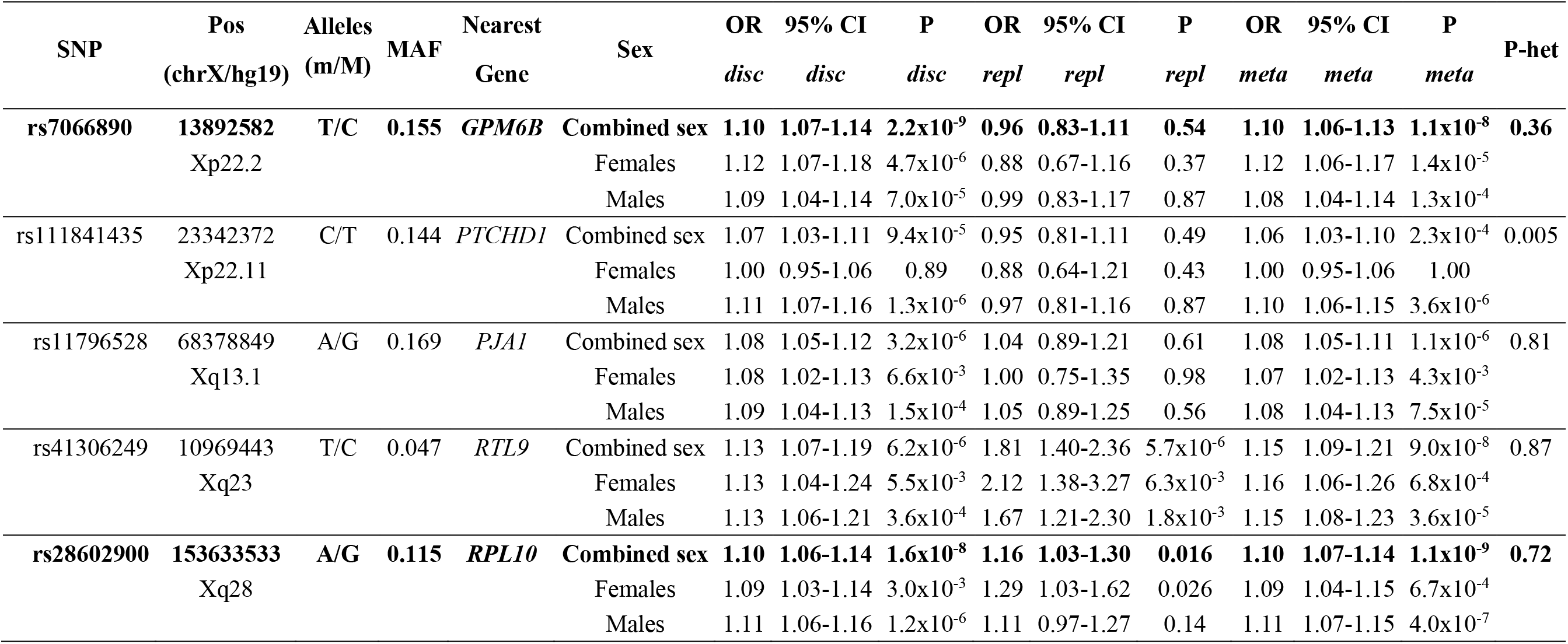
Lead SNPs in loci passing the suggestive threshold (P < 10^−5^) in the combined sex meta-analysis or single-sex meta-analyses. m/M: minor allele/major allele; MAF: minor allele frequency; P-het: P-value from the sex-heterogeneity test, ORs are calculated with respect to the minor allele. SNPs on lines highlighted in bold are genome-wide significant (P < 5×10^−8^). *disc*: discovery meta-analysis; *repl*: replication meta-analysis; *meta*: full meta-analysis.

### Independent PD replication meta-analysis

In two independent datasets, we tested the association with PD of the two lead SNPs from the genome-wide significant loci. The rs28602900 association with PD significantly replicated (OR = 1.16, p = 0.016), whereas rs7066890 was imputed in only one of the two datasets and showed a discordant direction of effect (OR = 0.96, p = 0.54). Thus, the rs7066890 association did not replicate but remained genome-wide significant after meta-analysis with the replication (p = 1.1×10^−8^, **Table 3**).

### Functional annotation via eQTL analysis

The identified loci lie in regions with multiple genes, so we sought to identify the potentially causal genes through eQTL analysis. To this aim, we queried the genome-wide significant SNPs in the GTEx v8 ^39^ and Braineac ^40^ databases, which both contain gene expression data from brain tissues and include eQTL analyses of X-chromosome SNPs.

All the following associations for rs28602900 were FDR significant in GTEx. rs28602900 was associated with decreased *RPL10* expression in 11 queried brain tissues. rs28602900 was also associated with decreased *PLXNA3* expression in 6 brain tissues (cerebellum, frontal cortex, cortex, putamen, cerebellar hemisphere, anterior cingulate cortex); increased *FAM50A* expression in 4 brain tissues (cerebellum, hypothalamus, cerebellar hemisphere, nucleus accumbens); increased *ATP6AP1* expression in the frontal cortex only; and decreased *DNASE1L1* expression in the cerebellum only. In Braineac, rs28602900 was significantly associated with decreased *RPL10* and *PLXNA3* average expression across brain tissues (p < 0.05, FDR-corrected). The rs28602900 association with *RPL10* was also FDR significant at the single tissue level in the cerebellum and temporal cortex tissues. No FDR-significant eQTL associations were observed in brain tissues for rs7066890 (*GPM6B* locus) in GTEx or Braineac.

The *RPL10* eQTL colocalized across 11 brain tissues with the locus associated with PD risk (PP4 between 0.90 and 0.97, **Fig S1**). The colocalization is illustrated for the putamen (**Fig 3A**). The *FAM50A* eQTLs in 4 brain tissues and the *ATP6AP1* eQTL in frontal cortex all also colocalized with the identified PD risk locus (PP4 between 0.90 and 0.95, and lower PP4 in other brain tissues, **Fig S1**). On the other hand, the *PLXNA3* eQTL association is driven by a nearby stronger *PLXNA3* eQTL, which is in low linkage disequilibrium with the locus associated with PD risk. This is illustrated for the putamen (PP4 = 0, **Fig 3B**) and occurred similarly in the 5 other brain tissues with a significant eQTL for *PLXNA3* (PP4 ≈ 0). Similarly, the *DNASE1L1* eQTL in the cerebellum was driven by a locus in low linkage disequilibrium with our PD risk association locus (PP4 = 0.4).

**Figure 3.**
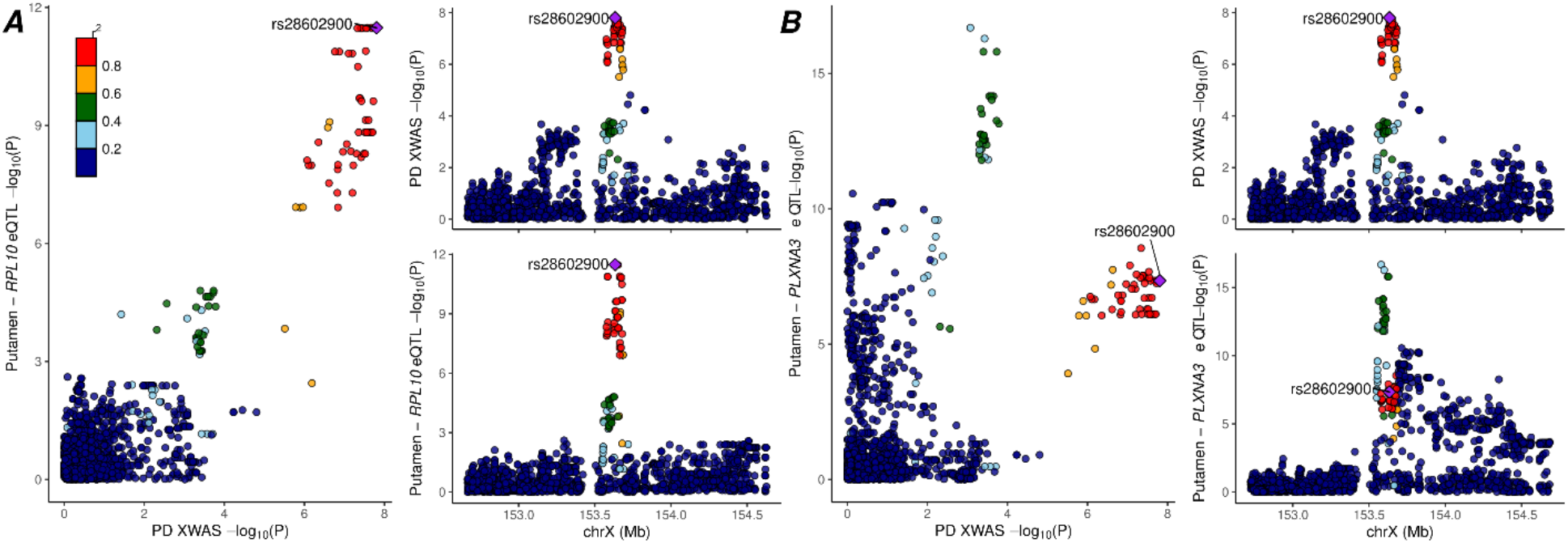
Colocalization analyses between association with PD risk and association with gene expression in brain tissues. **(A)** The locus, on Xq28, associated with PD risk colocalized with the *RPL10* eQTL in the putamen (and in the ten other GTEx brain tissues with FDR-significant *RPL10* eQTLs, data not shown). **(B)** The locus, on Xq28, associated with PD risk did not colocalize with the *PLXNA3* eQTL in the putamen (or in other brain tissues with FDR-significant *PLXNA3* eQTLs). Rather, the eQTL signal is driven by a nearby eQTL locus in low linkage disequilibrium with the PD risk locus.

### Association with putamen volume in the UK Biobank

To further assess and validate the role of the two genome-wide significant loci, we tested their association in UK Biobank with the putamen volume, an established PD biomarker ^45^. In a sample composed of 32,896 British individuals, excluding PD cases and PD-proxy-cases, the lead SNP in the *RPL10* locus, rs28602900, was significantly associated with decreased putamen volume (β= - 11.01, σ = 4.35, p = 0.011). The lead SNP in the *GPM6B* locus, rs7066890, was not significantly associated with putamen volume.

## Discussion

Our study demonstrates a genome-wide significant association of PD with two X-chromosome loci and paves the way for further XWAS studies to decipher the role of common X-chromosome variants in neurodegenerative diseases. This analysis expands our understanding of the genes involved in PD. These new loci will help refine polygenic risk scores to determine an individual’s genetic susceptibility to PD. We have also leveraged eQTL data to focus attention on a gene new to PD which may have important pathogenic implications.

The *RPL10* locus has the strongest supports across our analyses and was replicated in an independent PD meta-analysis. rs28602900, the top SNP in the locus, was significantly associated with increased risk in our combined sex meta-analysis and with reduced putamen volume in the UKB analysis. While this locus is in a high gene-density region, we were able to rank-order the list of potentially causal genes through functional annotations based on gene expression analysis. The most significant brain eQTLs from this locus point towards a regulation of *RPL10*, but *FAM50A, ATP6AP1* could also be putative candidates based on this evidence. The expression association signals were stronger for *RPL10* than for *FAM50A* or *ATP6AP1* and for *RPL10* reached FDR-corrected significance across all 11 brain tissues in GTEx. In addition, the expression analyses in Braineac also showed a significant association between rs28602900 and the average *RPL10* expression across all brain tissues (but no significant association for *FAM50A* or *ATP6AP1*). In both GTEx and Braineac, *RPL10* expression was lower in carriers of the rs28602900 minor allele. *RPL10*, ribosomal protein L10, has been associated with intellectual disability ^46^, cerebellar hypoplasia ^47^, microcephaly ^48^, and autism spectrum disorder ^49,50^. More generally, in a gene and protein network analysis of PD, Alzheimer’s disease and amyotrophic lateral sclerosis, Monti *et al*. emphasized that 6 out of 25 PD-specific proteins were ribosomal ^51^. Additionally, ribosomal proteins interact with *LRRK2* and elevate kinase activity ^52^. Specifically, the ribosomal protein, *RPS15*, was shown to be a key pathogenic *LRRK2* substrate mediating dopamine neuron degeneration ^52^. The reduced expression of several ribosomal proteins in PD brain tissues is associated with alpha-synuclein aggregation ^53^.

The second locus, near *GPM6B*, did not replicate in an independent PD meta-analysis and did not have any eQTL signals in brain tissue that survived FDR correction. The top SNP at the locus is intronic in *GPM6B*, whose associated protein is highly expressed in brain. The locus is also situated near a number of other brain-expressed proteins such as *OFD1, TRAPPC2*, and *GEMIN8*, but for the time being, the gene associated with PD risk at this locus remains uncertain.

Interestingly, a linkage analysis in multiplex PD families identified a large PD X-linked region, *PARK12*, spanning Xq21-q25 ^9^. Our analysis highlights a well-defined signal within this region, with a lead variant missense of *RTL9* (OR = 1.15, p = 9.0×10^−8^, **Table 3**). Linkage studies are well-powered to discover genetic loci associated with disease and reduce the risk of false positives due to population structure, but this approach often cannot fine-map the causal gene. Our complementary XWAS approach allowed us to refine this original signal and emphasized the most likely polymorphism increasing disease risk at this locus.

The main limitation of our study is that while we have three suggestive loci and two loci meeting the genome-wide significance threshold—including one replicating in an independent PD meta-analysis and strongly supported by eQTL and imaging analyses—we do not have access to additional replication datasets to validate more of these loci. Two large datasets used in the recent PD GWAS ^2^ could not, unfortunately, be used here because access to the data is restricted by the investigative teams. When additional datasets become available, future studies will allow further validation and extension of the findings presented here. In addition, the inclusion of UK Biobank proxy phenotypes that rely on participant reports of family history likely injects some diagnostic uncertainty into those data, but we believe this is outweighed by the gain in statistical power from being able to include this large cohort in our analyses.

In conclusion, our work provides compelling evidence for two novel loci on the X chromosome that increase risk for PD and that can be used to refine polygenic risk scores of PD. Most critically, these results focus attention on a new-to-PD gene, *RPL10*, that should further increase enthusiasm for the role of ribosomal proteins in PD pathogenesis ^51,52^.

## Data Availability

IPDGC NeuroX data available at: https://www.ncbi.nlm.nih.gov/projects/gap/cgi-bin/study.cgi?study_id=phs000918.v1.p1
NINDS data available at:
https://www.ncbi.nlm.nih.gov/projects/gap/cgi-bin/study.cgi?study_id=phs001172.v1.p2
APDGC data available at:
https://www.ncbi.nlm.nih.gov/projects/gap/cgi-bin/dataset.cgi?study_id=phs000394.v1.p1
NGRC data available at:
https://www.ncbi.nlm.nih.gov/projects/gap/cgi-bin/study.cgi?study_id=phs000196.v3.p1
EBI MERGE data available at:
https://www.ebi.ac.uk/ega/studies/EGAS00000000034
UK Biobank data available at:
http://biobank.ctsu.ox.ac.uk/crystal/
PDCGC data available at:
https://www.ncbi.nlm.nih.gov/projects/gap/cgi-bin/study.cgi?study_id=phs001664.v1.p1
ACT data available at:
https://www.niagads.org/datasets/ng00034
For McGill please contact the Quebec Parkinson Network:
http://rpq-qpn.ca/en/

https://www.ncbi.nlm.nih.gov/projects/gap/cgi-bin/study.cgi?study_id=phs001172.v1.p2

https://www.ncbi.nlm.nih.gov/projects/gap/cgi-bin/dataset.cgi?study_id=phs000394.v1.p1

https://www.ncbi.nlm.nih.gov/projects/gap/cgi-bin/study.cgi?study_id=phs000196.v3.p1

https://www.ebi.ac.uk/ega/studies/EGAS00000000034

http://biobank.ctsu.ox.ac.uk/crystal/

https://www.ncbi.nlm.nih.gov/projects/gap/cgi-bin/study.cgi?study_id=phs001664.v1.p1

https://www.niagads.org/datasets/ng00034

http://rpq-qpn.ca/en/

## Acknowledgments

Funding for this study was provided by the Iqbal Farrukh & Asad Jamal Center for Cognitive Health in Aging, and the NIH (AG060747 and AG047366). ZGO is supported by the Fonds de recherche du Québec - Santé (FRQS) Chercheurs-boursiers award, in collaboration with Parkinson Quebec, and by the Young Investigator Award by Parkinson Canada.

We would like to thank all of the subjects who donated their time and biological samples to be a part of this study.

## Acknowledgment for the use of PD datasets

PD data included the International Parkinson’s Disease Consortium (IPDGC), NeuroX dataset provided through dbGaP accession phs000918.v1.p1. We thank the IPDGC and the funding source 100064005/11348001. National Institute of Neurological Disorders and Stroke, National Institute on Aging, National Institutes of Health, Bethesda, MD, and United States Army Medical Research Acquisition Activity (USAMRAA), Fort Detrick, MD/Department of Defense, DC, USA. (PI Andrew Singleton, PhD, and Mike Nalls, PhD). PD data also included the National Institute of Neurological Disorders and Stroke (NINDS) Parkinson’s Disease dataset provided through dbGaP accession phs001172.v1.p2 (PI Andrew Singleton, PhD, and John Hardy, PhD). PD data also included the Autopsy-Confirmed Parkinson Disease GWAS Consortium (APDGC) dataset provided through dbGaP accession (phs000394.v1.p1) (PI Jeffery Vance, MD, PhD, Collaborators: Thomas Beach, MD, PhD, Karen Marder, MD, MPH, Tatiana M. Foroud, Ted M. Dawson, MD, PhD, Matthew P. Frosch, MD, PhD, Dennis W. Dickson, MD, Samuel M. Goldman, MD, MPH, Harry Vinters, MD, Deborah Mash, PhD, James Leverenz, MD, Thomas J. Montine, MD, PhD, John Q. Trojanowski, MD, PhD, Vivianna Van Deerlin, MD, PhD) with funding sources: P50 NS071674-01S1 NIH, X01 HG006076-01 NIH, P50 NS072187-01S2 NIH, and genotyping center Johns Hopkins University Center for Inherited Disease Research (CIDR), Baltimore, MD, USA (HHSN268201100011I NHGRI and NIH). PD data also included the Genome-Wide Association Study of Parkinson Disease: Genes and Environment dataset provided through dbGaP accession (phs000196.v3.p1) (PI Haydeh Payami, PhD, and co-investigators: John Nutt, MD, Cyrus Zabetian, MS, MD, Stewart Factor, DO, Eric Molho, MD, Donald Higgins, MD) with funding source: 5R01NS36960-10 and genotyping center Johns Hopkins University Center for Inherited Disease Research (CIDR), Baltimore, MD, USA (HHSN268200782096C NIH). PD data also included the Parkinson’s Disease Cognitive Genetics Consortium (PDCGC) Stage I, NeuroX Dataset (phs001664.v1.p1) for which we acknowledge the Pacific Northwest Udall Center (P50 NS062684). Healthy individuals from the Adult Change in Thought (ACT) study, which is a longitudinal prospective cohort study that began in 1994 (UO1 AG006781, UO1 HG004610, UO1 HG006375, U01 HG008657). PD data also included the dataset generated by the Wellcome Trust Case-Control Consortium 2 (WTCCC2) from UK patients with Parkinson’s disease and UK control individuals from the 1958 Birth Cohort and National Blood Service (available through the EBI, accession EGAD00000000022 and EGAD00000000057). This study makes use of data generated by the Wellcome Trust Case-Control Consortium. A full list of the investigators who contributed to the generation of the data is available from www.wtccc.org.uk. Funding for the project was provided by the Wellcome Trust under award 076113, 085475 and 090355. Part of the data used in the preparation of this article were obtained from the AMP PD Knowledge Platform. For up-to-date information on the study, https://www.amp-pd.org.AMP PD – a public-private partnership – is managed by the FNIH and funded by Celgene, GSK, the Michael J. Fox Foundation for Parkinson’s Research, the National Institute of Neurological Disorders and Stroke, Pfizer, Sanofi, and Verily. The AMP-AD data included clinical data and biosamples obtained from the Fox Investigation for New Discovery of Biomarkers (BioFIND), the Harvard Biomarker Study (HBS), the Parkinson’s Progression Markers Initiative (PPMI), and the Parkinson’s Disease Biomarkers Program (PDBP). BioFIND is sponsored by The Michael J. Fox Foundation for Parkinson’s Research (MJFF) with support from the National Institute for Neurological Disorders and Stroke (NINDS). The BioFIND Investigators have not participated in reviewing the data analysis or content of the manuscript. For up-to-date information on the study, visit michaeljfox.org/biofin. Harvard NeuroDiscovery Biomarker Study (HBS) is a collaboration of HBS investigators funded through philanthropy and NIH and Non-NIH funding sources. A full list of HBS investigator can found at https://neurodiscovery.harvard.edu/biomarkers-core. The HBS Investigators have not participated in reviewing the data analysis or content of the manuscript. Parkinson’s Progression Markers Initiative (PPMI) – a public-private partnership – is funded by the Michael J. Fox Foundation for Parkinson’s Research and funding partners, including Abbvie, Allergan, Amathus therapeutics, Avid radiopharmaceuticals, Biogen, BioLegend, Bristol-Myers Squibb, Celgene, Denali, GE Healthcare, Genentech (Roche group), GSK, Janssen Neuroscience (Johnson-Johnson group), Lilly, Lundbeck, Merck, MSD, Pfizer, Piramal, Prevail, Roche, Sanofi Genzyme, Servier, Takeda, Teva, UCB, Verily, Voyager Therapeutics, Golub Capital. The PPMI, Investigators have not participated in reviewing the data analysis or content of the manuscript. For up-to-date information on the study, visit www.ppmi-info.org. Parkinson’s Disease Biomarker Program (PDBP) consortium is supported by the NINDS at the National Institutes of Health. A full list of PDBP investigators can be found at https://pdbp.ninds.nih.gov/policy. The PDBP Investigators have not participated in reviewing the data analysis or content of the manuscript. The McGill cohort was financially supported by grants from the Michael J. Fox Foundation, the Canadian Consortium on Neurodegeneration in Aging (CCNA), the Canada First Research Excellence Fund (CFREF), awarded to McGill University for the Healthy Brains for Healthy Lives initiative (HBHL), and Parkinson Canada. The access to part of the participants for this research has been made possible thanks to the Quebec Parkinson’s Network (http://rpq-qpn.ca/en/).

## Acknowledgement for the use of the UK Biobank data

This research has been conducted using the UK Biobank Resource under Application Number 45420.

## Author contributions

Y.L.G processed and analyzed data, designed analyses, designed study, and wrote paper. V.N. processed data, designed study, and supervised work. M.E.B., S.J.E., and G.K. processed data. E.Y., L.K., J.A.R., and Z.G.O acquired and processed data. M.D.G designed study, supervised work, and wrote paper. Y.L.G and V.N. contributed equally to this work.

