## Supplementary Materials for "Common X-chromosome variants are associated with Parkinson’s disease risk"

**Supplementary Figures**

**Figure S1. Posterior probability of colocalization (PP4) between association with PD risk and gene expression at the *RPL10* locus, considering all genes within +/- 1MB from the lead SNPs in 12 GTEx brain tissues.**

**Figure S2. Posterior probability of colocalization (PP4) between association with PD risk and gene expression at the *GPM6B* locus, considering all genes within +/- 1MB from the lead SNPs in 12 GTEx brain tissues.**

**Figure S3. Posterior probability of colocalization (PP4) between association with PD risk and gene expression at the *PJA1* locus, considering all genes within +/- 1MB from the lead SNPs in 12 GTEx brain tissues.**

**Figure S4. Posterior probability of colocalization between association with PD risk and gene expression at the *PTCHD1* locus, considering all genes within +/- 1MB from the lead SNPs in 12 GTEx brain tissues.**

**Supplementary Tables**

**Table S1. Autosomal variants excluded per quality control step prior to ancestry determination and genetic principal component computation.**

**Table S2. Number of individuals per ancestry per cohort.**

**Table S3. X chromosome variants excluded per quality control step prior to imputation.**

**Table S4. Number of Northwestern Europeans excluded for genotype missingness, sex discrepancy and relatedness in each cohort.**

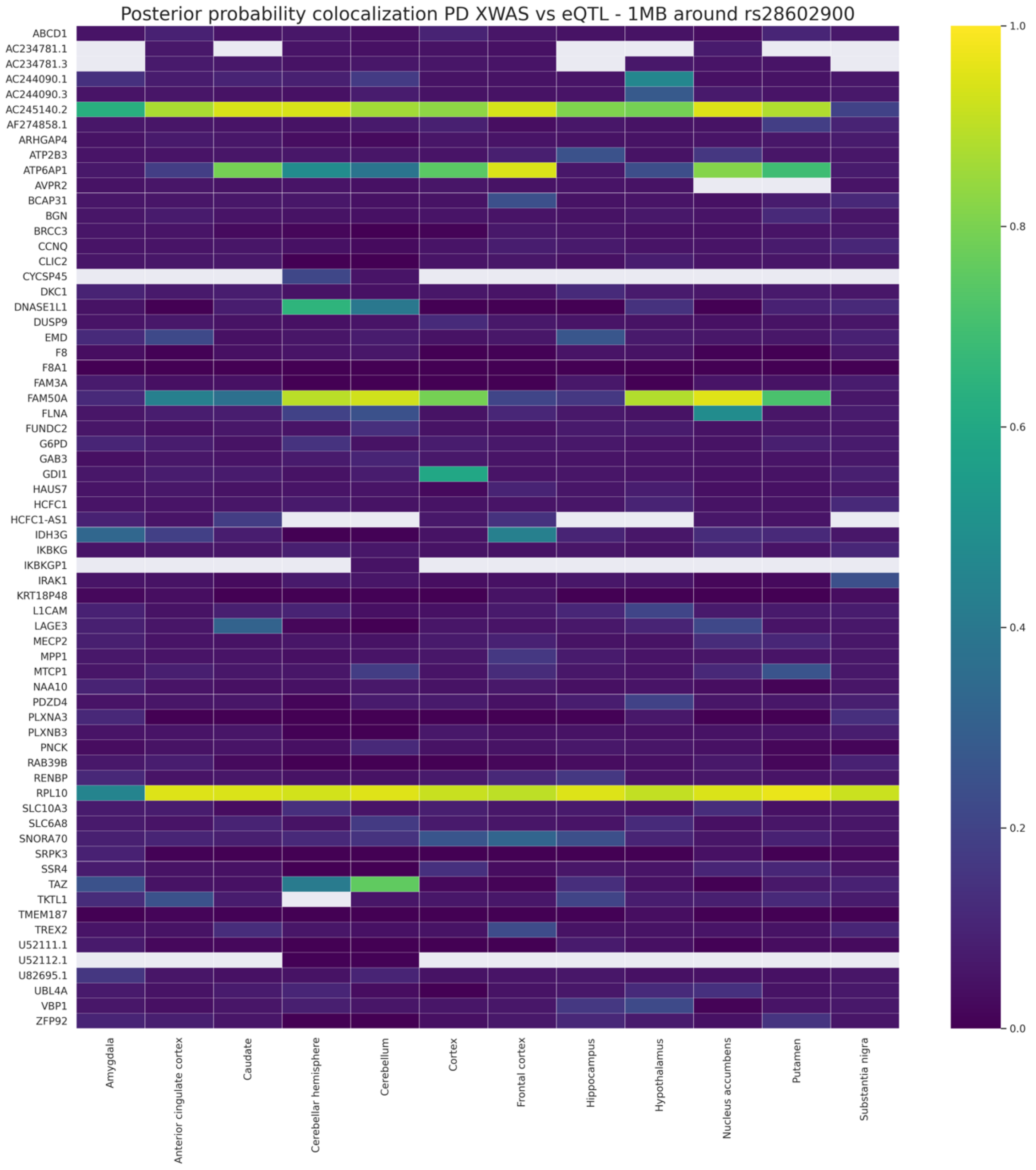


**Figure S1. Posterior probability of colocalization (PP4) between association with PD risk and gene expression at the *RPL10* locus, considering all genes within +/- 1MB from the lead SNPs in 12 GTEx brain tissues.**

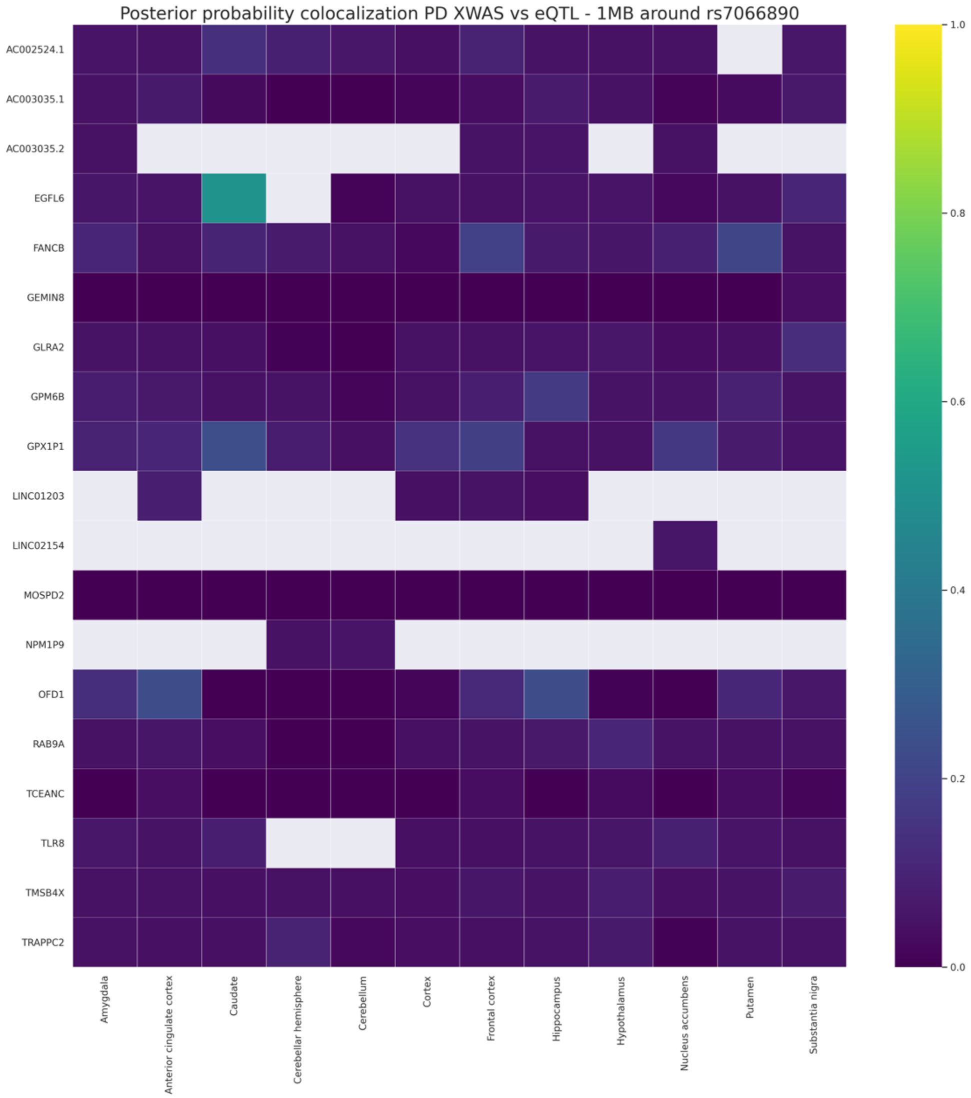


**Figure S2. Posterior probability of colocalization (PP4) between association with PD risk and gene expression at the *GPM6B* locus, considering all genes within +/- 1MB from the lead SNPs in 12 GTEx brain tissues.**

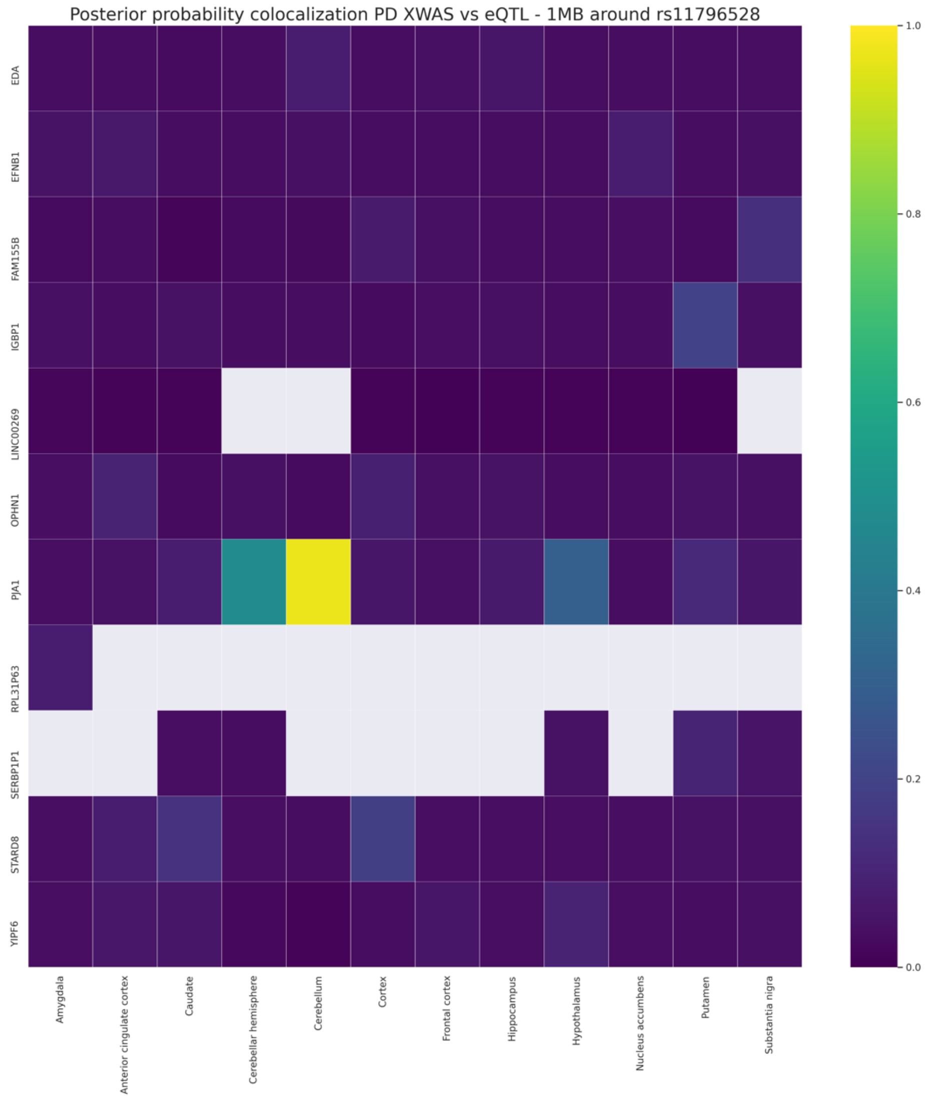


**Figure S3. Posterior probability of colocalization (PP4) between association with PD risk and gene expression at the *PJA1* locus, considering all genes within +/- 1MB from the lead SNPs in 12 GTEx brain tissues.**

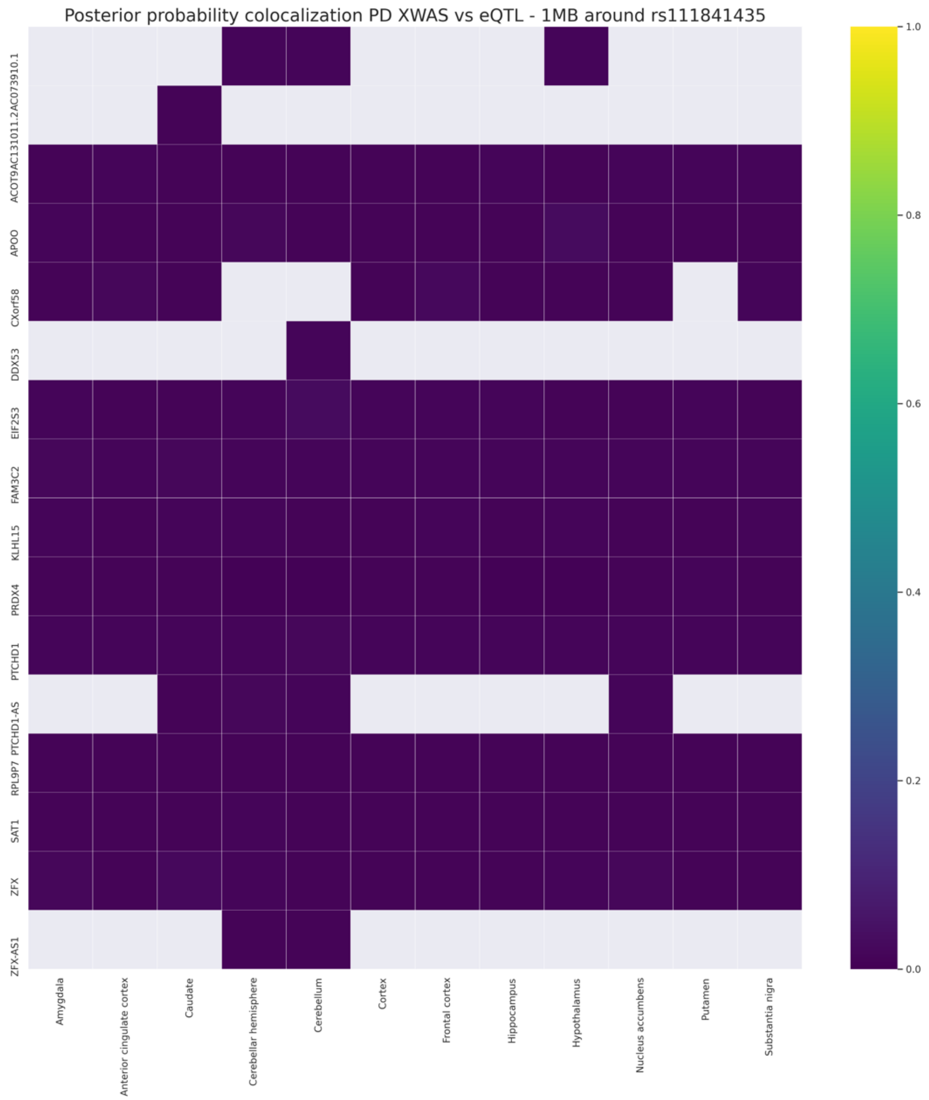


**Figure S4. Posterior probability of colocalization between association with PD risk and gene expression at the *PTCHD1* locus, considering all genes within +/- 1MB from the lead SNPs in 12 GTEx brain tissues.**

**Table S1**. **Autosomal variants excluded per quality control step prior to ancestry determination and genetic principal component computation.** (see Methods) McGill autosomes were processed separately.

| Cohort | Autosomes hg19 | Multi-allelic | CNV and Segmental duplication regions | Duplicate vars | Primer Polymorphism | Monomorphic | Wrong alleles | Check with HRC reference panel | Genotyping rate | Individuals excluded due to missingness | Hardy-Weinberg equilibrium | Autosomal SNPs after QC |
|---|---|---|---|---|---|---|---|---|---|---|---|---|
| **APDGC** | 965259 | 0 | 415 | 0 | 4715 | 27718 | 13138 | 18330 | 5871 | 0 | 382 | 894960 |
| **IPDGC** | 203902 | 0 | 48777 | 0 | 55 | 4841 | 0 | 32326 | 1531 | 2 | 310 | 116062 |
| **NGRC** | 979887 | 10 | 448 | 4 | 4813 | 25163 | 26947 | 14423 | 954 | 0 | 2515 | 904610 |
| **NINDS** | 531666 | 182 | 2022 | 2 | 514 | 682 | 673 | 3355 | 4654 | 1 | 109 | 519473 |
| **EGAD22_illumina** | 491680 | 0 | 973 | 0 | 460 | 0 | 0 | 11467 | 958 | 16 | 3405 | 474417 |
| **EGAD57_illumina** | 579626 | 0 | 1976 | 0 | 807 | 6755 | 0 | 11742 | 2975 | 14 | 0 | 555371 |
| **EBI MERGE** | 765669 | 0 | 0 | 4 | 0 | 0 | 0 | 0 | 494935* | 39 | 1719 | 269011 |
| **ACT** | 546428 | 0 | 17772 | 0 | 546 | 266 | 28 | 2049 | 191 | 0 | 242 | 525334 |
| **PDCGC** | 201093 | 70 | 22 | 0 | 107 | 99347 | 5544 | 13800 | 525 | 3 | 342 | 81336 |
| **PDCGC & ACT** | 597731 | 0 | 0 | 0 | 0 | 3 | 0 | 0 | 588539** | 3 | 0 | 9189 |

* most of these variants were not overlapping between the two Illumina arrays in EGAD22 and EGAD57, and were thus removed after merge
** most of these variants were not overlapping between the two Illumina arrays in ACT and PDCGC, and were thus removed after merge

**Table S2. Number of individuals per ancestry per cohort.** By applying an ancestry percentage cut-off > 75%, the samples were stratified into the five super populations, South-Asians (SAS), East-Asians (EAS), Americans (AMR), Africans (AFR) and Europeans (EUR). Individuals who do meet the 75% cut-off were classified as ADMIX. Since most of the samples belonged to the European population, we also determined their percent ancestry to three major ethnicities, Northwestern (NWE), Southeastern (SEE), and Ashkenazi Jewish (AJE), using reference populations available from SNPweights v.2.1. European subjects were stratified into the above-mentioned ethnicities when their ancestry percentage was greater than 50%. Europeans who do not meet the 50% cut-off were classified as EUR. Note that NWE, SEE, AJE individuals are also EUR but are only counted in their respective column.

| Cohort | NWE | SEE | AJW | EUR | ADMIX | AFR | AMR | SAS | EAS |
|---|---|---|---|---|---|---|---|---|---|
| **AMP PD WGS** | 2636 | 254 | 709 | 163 | 97 | 27 | 24 | 6 | 25 |
| **APDGC** | 760 | 39 | 117 | 17 | 19 | 16 | 2 | 0 | 7 |
| **EBI MERGE** | 4937 | 32 | 48 | 26 | 27 | 4 | 1 | 10 | 2 |
| **IPDGC** | 7940 | 2175 | 389 | 323 | 573 | 0 | 0 | 0 | 0 |
| **NGRC** | 3651 | 136 | 130 | 53 | 15 | 0 | 1 | 0 | 0 |
| **NINDS** | 1469 | 88 | 99 | 39 | 39 | 0 | 6 | 0 | 0 |
| **PDCGC & ACT MERGE** | 2410 | 138 | 135 | 60 | 238 | 37 | 27 | 1 | 59 |
| **McGill** | 1994 | 142 | 388 | 73 | 63 | 19 | 14 | 4 | 13 |

**Table S3**. **X chromosome variants excluded per quality control step prior to imputation.** PAR: pseudo-autosomal regions on the X-chromosome. (See Methods for more details)

| Cohort | X hg19 | Multi-allelic | Primer Polymorphism | Wrong alleles | PAR | Genotyping rate | Differential missingness cases vs controls | HWE in females | Monomorphic | Differential missingness males vs females | Differential MAF males vs females | Check with HRC reference panel | N SNPs prior to imputation |
|---|---|---|---|---|---|---|---|---|---|---|---|---|---|
| **APDGC** | 21825 | 0 | 1986 | 325 | 0 | 10 | 0 | 0 | 1801 | 2 | 0 | 205 | 17496 |
| **IPDGC** | 4229 | 0 | 38 | 1311 | 55 | 52 | 41 | 0 | 444 | 125 | 1 | 498 | 1664 |
| **NGRC** | 23109 | 0 | 2118 | 369 | 0 | 1 | 0 | 0 | 542 | 120 | 0 | 171 | 19788 |
| **NINDS** | 13203 | 2 | 754 | 58 | 0 | 356 | 90 | 0 | 77 | 60 | 1 | 73 | 11732 |
| **EGAD22_illumina** | 14480 | 0 | 731 | 0 | 0 | 67 | 0 | 26 | 14 | 401 | 6 | 280 | 12955 |
| **EGAD57_illumina** | 14189 | 0 | 858 | 0 | 0 | 1286 | 0 | 0 | 27 | 101 | 20 | 70 | 11827 |
| **EBI MERGE** | 19247 | 0 | 0 | 0 | 0 | 13368* | 104 | 0 | 0 | 18 | 0 | 30 | 5727 |
| **ACT** | 8939 | 0 | 0 | 0 | 0 | 0 | 0 | 0 | 31 | 0 | 0 | 0 | 8908 |
| **PDCGC** | 4210 | 0 | 37 | 2320 | 42 | 13 | 0 | 2 | 209 | 6 | 1 | 258 | 1322 |
| **McGill** | 17942 | 0 | 746 | 0 | 1 | 142 | 278 | 82 | 690 | 145 | 1 | 90 | 15912 |

* these variants were not overlapping between the two Illumina arrays in EGAD22 and EGAD57

**Table S4. Number of Northwestern Europeans excluded for genotype missingness, sex discrepancy and relatedness in each cohort.**

| Cohort | NWE | Genotype missingness | Sex discrepancy | Relatedness | NWE after QC |
|---|---|---|---|---|---|
| **AMP PD WGS** | 2636 | 0 | 0 | 219 | 2417 |
| **APDGC** | 760 | 0 | 0 | 0 | 760 |
| **IPDGC** | 7940 | 27 | 1 | 238 | 7674 |
| **EBI MERGE** | 4937 | 0 | 29 | 309 | 4599 |
| **NGRC** | 3651 | 0 | 1 | 25 | 3625 |
| **NINDS** | 1469 | 0 | 0 | 28 | 1441 |
| **PDCGC & ACT** | 2410 | 0 | 2 | 168 | 2240 |
| **McGill** | 1994 | 48 | 65 | 90 | 1791 |
